# Trends of enterovirus D68 concentrations in wastewater in two California communities parallel trends of statewide confirmed cases, February 2021 – April 2023

**DOI:** 10.1101/2023.08.24.23294571

**Authors:** Alexandria B. Boehm, Debra A. Wadford, Bridgette Hughes, Dorothea Duong, Alice Chen, Tasha Padilla, Chelsea Wright, Lisa Moua, Teal Bullick, Maria Salas, Christina Morales, Bradley J. White, Carol A. Glaser, Duc J. Vugia, Alexander T. Yu, Marlene K Wolfe

## Abstract

In this retrospective study, enterovirus D68 (EVD68) genomic RNA in wastewater solids was measured longitudinally at two California wastewater treatment plants twice per week for 26 months. EVD68 RNA was undetectable except when concentrations increased between mid-July and mid-December 2022, during which time EVD68 cases were confirmed in the state.

## Background

Enterovirus D68 (EVD68) was first recognized as a respiratory virus in 1962^1^. In 2014, unprecedented large outbreaks of EVD68 associated with severe respiratory illnesses occurred in children in the United States (US)^2^, which coincided with a subsequent rise in cases of acute flaccid myelitis (AFM)^3^. The US Centers for Disease Control and Prevention (CDC) began active AFM monitoring in 2014, and as of July 2023, there have been 729 confirmed cases^4^. Most patients develop AFM between August and November. Cyclical peaks in AFM incidence were observed every two years (2014, 2016, and 2018) prior to the COVID-19 pandemic.

CDC began active sentinel surveillance for EVD68 in 2017 through the New Vaccine Surveillance Network and detected a greater number of EVD68 cases during July-August 2022^5^. However, there is no formal state or local EVD68 surveillance. With the emergence of wastewater surveillance (WWS) as a method to monitor pathogens at various population levels^6^, we sought to assess the feasibility of applying WWS to better understand EVD68 circulation. WWS could provide an early alert system for public health authorities to mitigate possible increases in severe acute respiratory illness and AFM, and to enhance physician awareness during times of increased circulation. Two recent studies reported EVD68 RNA in wastewater in 2021 and concordance with confirmed infections in Israel and UK^7,8^. In this retrospective, longitudinal study, we developed an EVD68 assay, deployed it for WWS and compared WWS with laboratory-confirmed EVD68 cases.

### This Study

We tested the EVD68-specific primers and probe developed by Wylie et al^9^ for their sensitivity and specificity *in silico*, and *in vitro* against virus panels, intact viruses, and cDNA gene blocks (see Appendix for in vitro testing details). *In silico* and *in vitro* testing indicated no cross reactivity with non-EVD68 sequences deposited in National Center for Biotechnology Information (NCBI) and non-target viral gRNA, respectively. However, the assay did not detect the EVD68 variant circulating in fall 2022 in the Northern Hemisphere (EVD68-2022). We therefore developed a new set of primers, and modified the probe, to amplify and detect in EVD68-2022 the same region of the polyprotein region (VP1) gene targeted by the Wylie assay. EVD68-2022 genome sequences were downloaded from NCBI in September 2022, aligned to identify conserved regions in VP1, and primers and probes were developed *in silico* (Table A1). New primers and probes were confirmed to be specific and sensitive *in silico* and *in vitro*. The EVD68 assay used in this study includes two forward and two reverse primers in order to ensure detection of all EVD68 variants (Table 1).

**Table 1.**
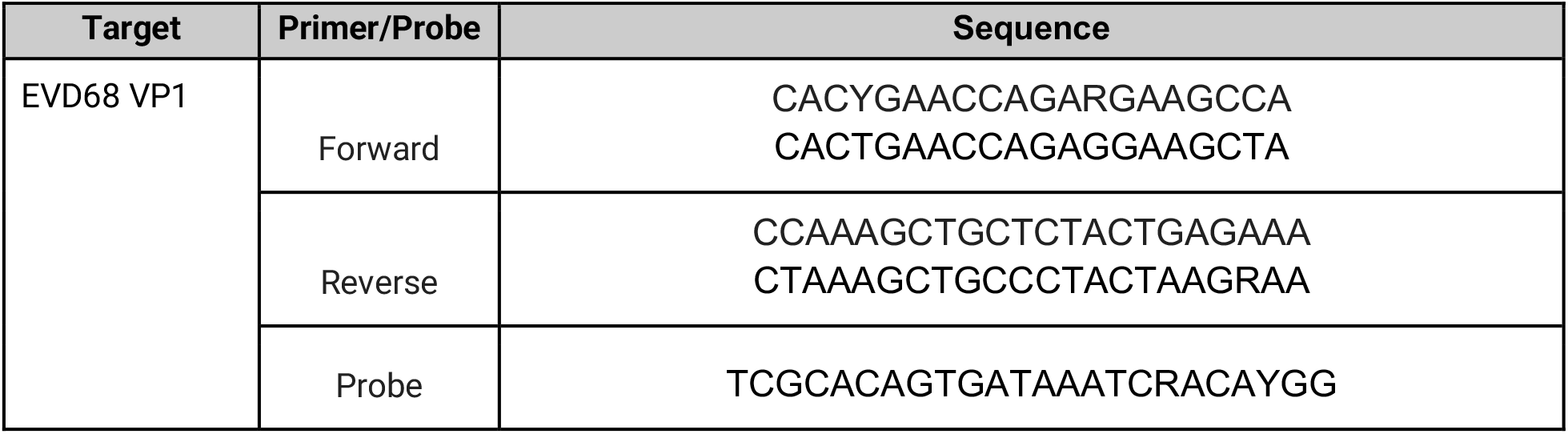
Forward and reverse primers and probe used in this study.

Wastewater solids samples were retrospectively selected from biobanked samples collected as part of a prospective WWS program. Samples were selected from San Jose (SJ) and Oceanside (OSP) wastewater treatment plants, serving 1.5 million people in Santa Clara County, CA, and 250,000 people in San Francisco County, CA, respectively (Figure A1). Two samples from each site per week collected between 2/1/21 and 4/14/23 were analyzed. Wastewater solids were thawed overnight; nucleic acids were extracted from 10 replicate sample aliquots as described elsewhere^10^. RNA was used as template in 10 replicate digital droplet RT-PCR wells for each sample to measure EVD68 RNA. We measured pepper mild mottle virus (PMMoV) RNA as an endogenous control. Details of RT-PCR chemistry and data processing are provided in the Appendix. Results are reported as copies per gram dry weight of solids (cp/g) and errors are 68% confidence intervals. The lower limit of detection is approximately 500 cp/g.

Between February 2021 and April 2023, EVD68 RNA concentrations in wastewater solids were non-detectable except for samples collected between mid-July and mid-December 2022. During this time, EVD68 RNA concentrations increased to approximately 10^5^ cp/g in mid-October at both plants (Figure 1a-b). Plots of EVD68 normalized by PMMoV are similar (Figure A2a-b).

**Figure 1.**
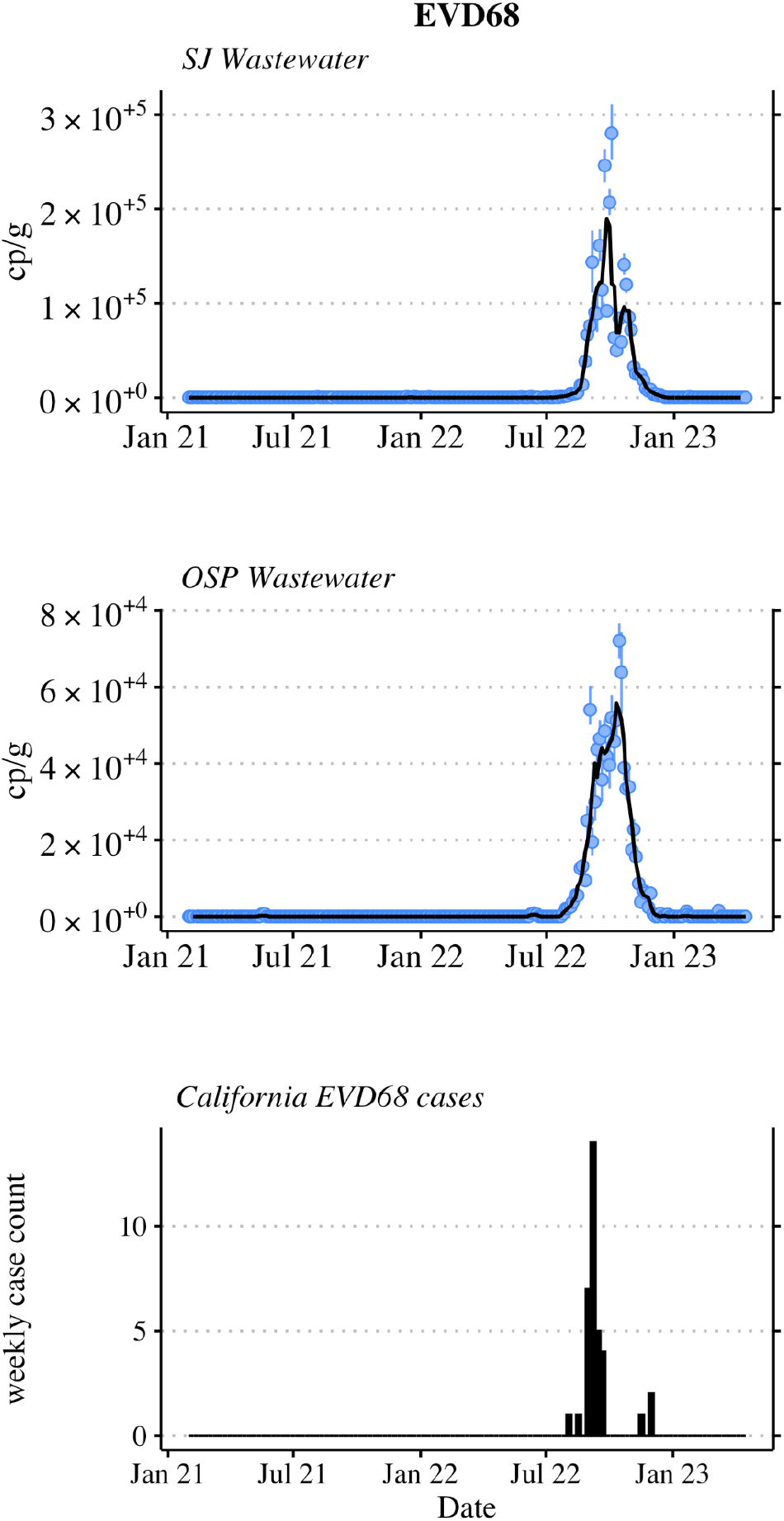
EVD68 RNA concentrations in wastewater solids at a. San Jose (SJ) and b. Oceanside (OSP) wastewater treatment plants, and c. weekly- and state-aggregated laboratory-confirmed EVD68 cases (bottom). Error bars on wastewater measurements represent 68% confidence intervals. Black lines on wastewater data represent five-sample trimmed means.

Although there is no active EVD68 surveillance in California, CDPH Viral and Rickettsial Disease Laboratory (VRDL) accepts EV-reactive specimens from hospitals, clinics, and local public health laboratories for strain typing. EV typing allows us to know, at a limited level, which EV types are circulating and associated with illness. Since 2020, CDPH VRDL has maintained an AFM surveillance program with CDC^11^. This enhanced AFM surveillance captures patient information but is not specific for EVD68, per se. However, EV testing and typing are conducted when warranted. The thirty-five EVD68 samples confirmed by VRDL from January 2021 through April 2023 are shown aggregated by week (Figure 1c).

Trends in EVD68 RNA wastewater concentrations at the two California communities match the trend in the state-aggregate, weekly-aggregated laboratory-confirmed EVD68 cases (Figure 1). Weekly median wastewater EVD68 RNA concentrations were positively correlated to weekly case counts (Kendall’s tau=0.47 and 0.50 for SJ and OSP respectively, p<0.001).

This study was reviewed by the State of California Health and Human Services Agency Committee for the Protection of Human Subjects and determined to be Exempt from oversight.

## Conclusions

Detection of EVD68 in wastewater over 26 months in two California communities corresponded strikingly with the trend of statewide laboratory-confirmed EVD68 cases. Passive EV surveillance and sentinel EVD68 surveillance in the US do not detect the majority of EV cases and focus primarily on the most severe EV cases causing acute respiratory illness, meningitis, and AFM. Most individuals infected with EV are either asymptomatic or have mild symptoms and would not likely be tested. For those who are tested, EV-specific testing is rare and a presumptive diagnosis is often based on the rhinovirus/enterovirus test on a respiratory virus panel assay without further characterization. Even in cases where EVD68 is identified, EVD68 infection is not a reportable disease and not necessarily captured in public health surveillance systems. During July-December 2022 when EVD68 RNA was detected and peaked in wastewater, case data were sparse, even when aggregated at the state level, as shown in Figure 1c.

Given that known recent EVD68 surges have been associated with severe pediatric respiratory illness and coincided with increased numbers of AFM cases, there is a benefit to enhancing EVD68 surveillance so that its circulation dynamics are better understood. According to the data reported herein, WWS can be applicable for this purpose. Because results from WWS are available 24 hours after sample collection, early warning about increases in circulating EVD68 can be provided to communities being monitored, irrespective of clinical testing. WWS for EVD68 can inform public health action, including when to issue alerts to clinicians of the potential for severe respiratory illnesses and AFM cases. Clinician awareness is critical for effective patient care because early recognition of severe cases, including AFM, will prevent unnecessary testing for other etiologies, optimize patient management, and may improve outcomes^11^. Our findings suggest such surveillance is possible through routine EVD68 monitoring of wastewater.

## Data Availability

Wastewater data are publicly available at https://doi.org/10.25740/qt551tn4819.

## Acknowledgments

We acknowledge Hugo Guevara and Chao-Yang Pan for their contributions to the EVD68 case surveillance and Vikram Chan-Herur for his support on assay design. B. White, D. Duong, and B. Hughes are employees of Verily Life Sciences; other authors declare no competing interests.

## Notes

### Funding Statement

This study was funded by a gift from the Sergey Brin Family Foundation to A. Boehm.

