## Appendix for "Trends of enterovirus D68 concentrations in wastewater in two California communities parallel trends of statewide confirmed cases, February 2021 – April 2023"

**Additional methods for EVD68 assay validation.** EVD68 assays were screened for specificity and sensitivity *in silico*, and *in vitro* against virus panels (NATRVP2.1-BIO and NATEVP-C from Zeptomatrix, Buffalo, NY), intact viruses (American Type Culture Collection (ATCC) VR-1826DQ and ATCC VR-1823D, Manassas, VA), and cDNA gene blocks. Primers, probes, and gene blocks were purchased from Integrated DNA Technologies (Coralville, IA).

For EVD68 assay *in vitro* sensitivity and specificity testing, nucleic acids were extracted from intact viruses using Chemagic Viral DNA/RNA 300 Kit H96 for Chemagic 360 (PerkinElmer, Waltham, MA). Nucleic acids were used undiluted as template in digital droplet (RT-)PCR singleton assays for sensitivity and specificity testing in single wells. The concentration of targets used in the in vitro specificity testing was between $10^3$ and $10^4$ copies per well. Negative (RT-)PCR controls were included on each plate.

**Additional pre-analytical methods for wastewater samples.** Wastewater solids were thawed overnight; nucleic acids were extracted from the 10 replicate sample aliquots using protocols described elsewhere[1]. Those protocols include suspending the solids in a buffer at a concentration of 75 mg/ml and using an inhibitor removal kit; together these processes alleviate potential inhibition while maintaining good assay sensitivity[1,2]. Nucleic-acids were obtained from 10 replicate sample aliquots. Each replicate nucleic-acid extract from each sample was subsequently stored between 8 and 273 days (median 266 days) for SJ and between 1 and 8 days (median 5 days) for OSP at -80°C and subjected to a single freeze thaw cycle. After the nucleic-acids were thawed, they were used immediately as template in the RT-PCR assays.

**Details of the PCR reactions.** The digital droplet (dd)RT-PCR methods applied to wastewater solids to measure PMMoV in a singleplex reaction are provided in detail elsewhere[3]. The EVD68 assay was run in multiplex using a probe-mixing approach and unique fluorescent molecules (indicated in parentheses) with other assays for which results are not described herein. The RT-PCR included primers and probes for EVD68 (FAM), hepatitis A virus (Cy5), pan-enterovirus (Cy5.5), and human norovirus GI (ATTO590). Primers and probes for assays were purchased from Integrated DNA Technologies (IDT, San Diego, CA).

Each 96-well PCR plate of wastewater samples included PCR positive controls for each target assayed on the plate in one well, PCR negative no template controls in two wells, and extraction negative controls (consisting of water and lysis buffer) in two wells. PCR positive controls consisted of viral gRNA or gene blocks.

ddRT-PCR was performed on 20 µl samples from a 22 µl reaction volume, prepared using 5.5 µl template, mixed with 5.5 µl of One-Step RT-ddPCR Advanced Kit for Probes (Bio-Rad 1863021), 2.2 µl of 200 U/µl Reverse Transcriptase, 1.1 µl of 300 mM dithiothreitol (DDT) and primer and probe mixtures at a final concentration of 900 nM (primers) and 250 nM (probe), respectively. EVD68 was measured in reactions using undiluted template whereas PMMoV was measured using template diluted 1:100 in molecular grade water.

Droplets were generated using the AutoDG Automated Droplet Generator (Bio-Rad, Hercules, CA). PCR was performed using Mastercycler Pro (Eppendforf, Enfield, CT) with the following cycling conditions: reverse transcription at 50°C for 60 minutes, enzyme activation at 95°C for 5 minutes, 40 cycles of denaturation at 95°C for 30 seconds and annealing and extension at 59°C (for EVD68) or 56°C (for PMMoV) for 30 seconds, enzyme deactivation at 98°C for 10 minutes then an indefinite hold at 4°C. The ramp rate for temperature changes were set to 2°C/second and the final hold at 4°C was performed for a minimum of 30 minutes to allow the droplets to stabilize. Droplets were analyzed using the QX200 or the QX600 Droplet Reader (Bio-Rad). A well had to have over 10,000 droplets for inclusion in the analysis. All liquid transfers were performed using the Agilent Bravo (Agilent Technologies, Santa Clara, CA).

Thresholding was done using QuantaSoft™ Analysis Pro Software (Bio-Rad, version 1.0.596) and QX Manager Software (Bio-Rad, version 2.0). Replicate wells were merged for analysis of each sample. In order for a sample to be recorded as positive, it had to have at least 3 positive droplets.

Concentrations of RNA targets were converted to concentrations per dry weight of solids in units of copies (cp)/g dry weight using dimensional analysis. The total error is reported as standard deviations and includes the errors associated with the Poisson distribution and the variability among the 10 replicates. Three positive droplets across 10 merged wells corresponds to a concentration between ~500 cp/g. Wastewater data are publicly available at https://doi.org/10.25740/qt551tn4819.

**Quality assurance and control.** Results are reported as suggested in the Environmental Microbiology Minimal Information (EMMI) guidelines[4] (Figure A3). Extraction and PCR negative and positive controls performed as expected (negative and positive, respectively). PMMoV measurements were used to assess potential for gross extraction failures as it is present in very high concentrations in the samples naturally, and lack of its detection, or abnormally low measurements might indicate gross extraction failures. The median (interquartile range) $\log_{10}$-transformed PMMoV was 9.1 (9.0-9.2) and 8.7 (8.6-8.8) $\log_{10}$ copies/g at SJ and OSP, respectively. The lowest measurements at the two sites were 8.5 (SJ) and 8.1 (OSP) $\log_{10}$ copies/g and given these lowest values are within an order of magnitude of the medians, we concluded that there was no gross extraction failures.

Additional details related to the EMMI guidelines are reported here. Thirty-six samples were selected at random for this analysis; this represents 8% of the samples processed in the study. The average (standard deviation) number of partitions (droplets) for each of the 2 reactions (across the 10 replicates) was 164,164 (38,851) for the reaction for PMMoV and 185,497 (24,410) for the reaction for EVD68. The volume of the partitions as reported by the machine vendor is 0.00085 µL. The mean and standard deviation of copies per partition for each target is shown in Table A2.

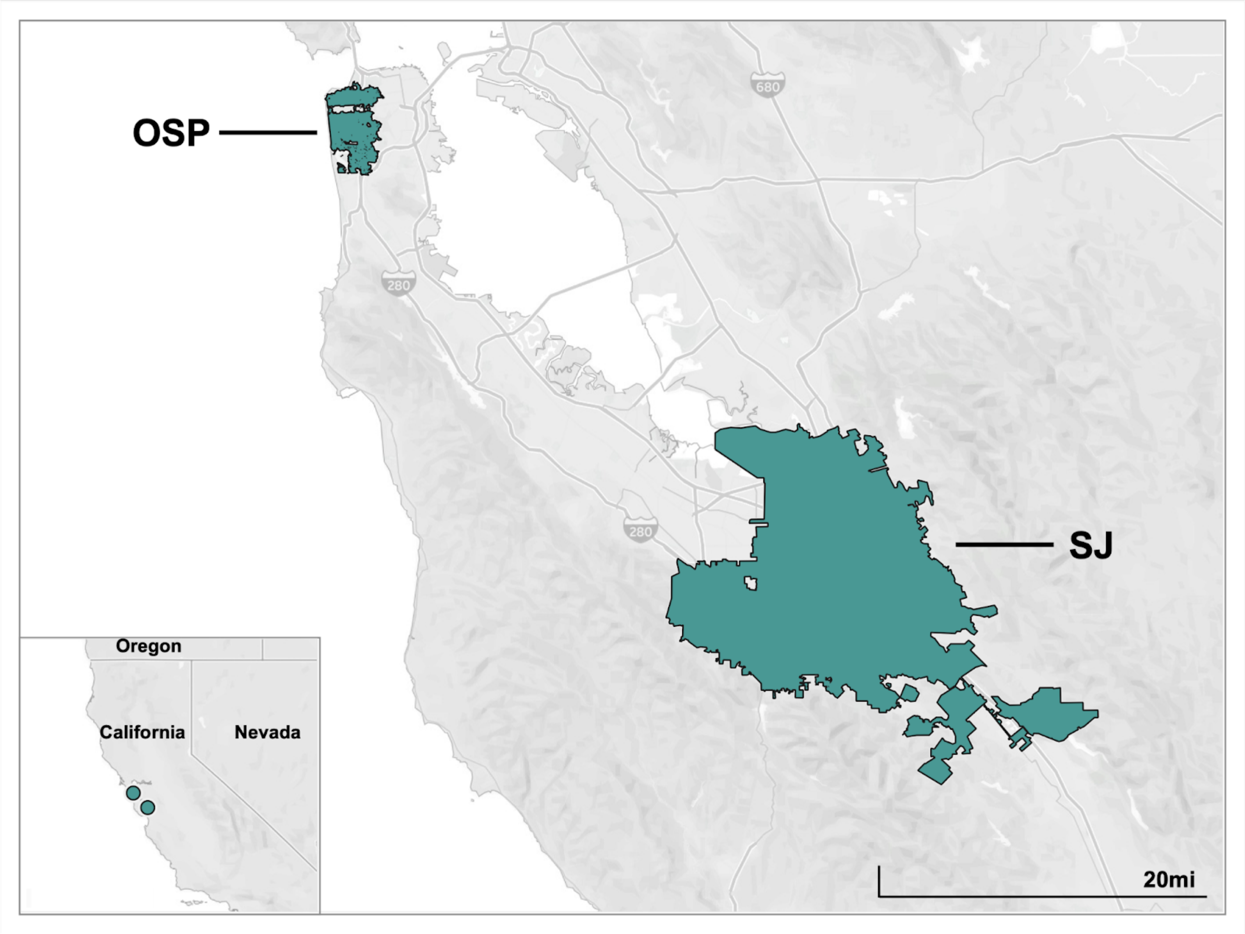


Appendix Figure A1. Map showing service areas for the two wastewater treatment plants in this study.

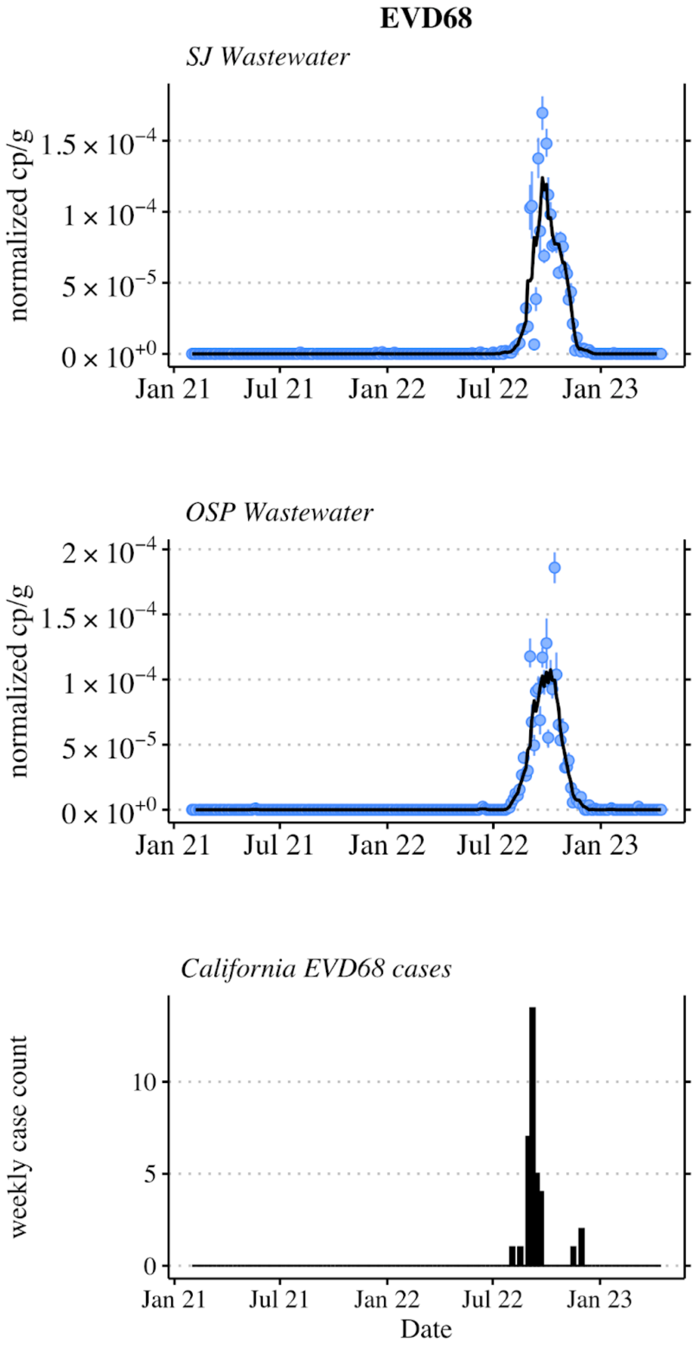


Figure A2. Concentrations of EVD68 RNA in wastewater solids at SJ and OSP normalized by concentrations of PMMoV (top two panels). Error bars show standard deviation of the ratio propagated by assuming the errors are symmetric and described by the larger of the lower or error bar from the original measurements. The black lines indicate 5-adjacent sample trimmed averages. Bottom panel shows the total number of laboratory-confirmed EVD68 cases each week aggregated across the state of CA.

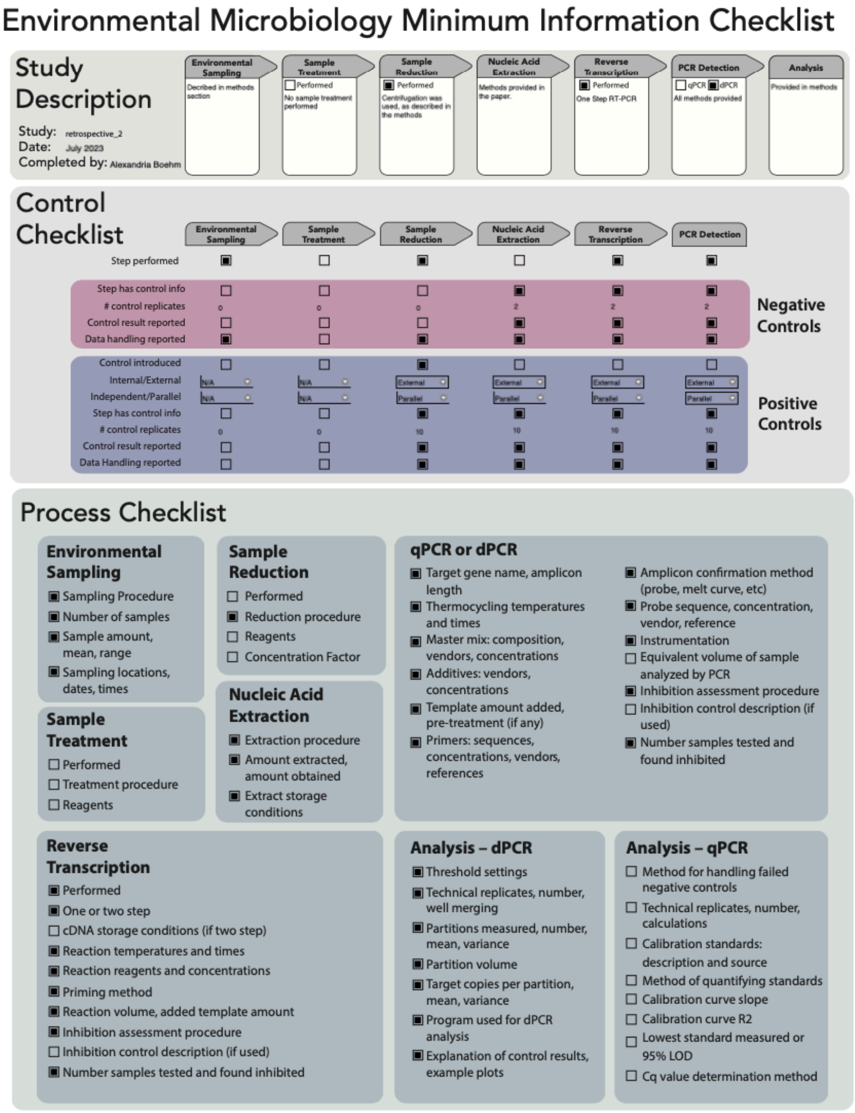


Figure A3. EMMI[4] checklist.

Table A1. Parameters used in the development of new primers and probes (internal oligo) using Primer3Plus (https://primer3plus.com/, accessed 7/16/23).

- Product size ranges: 60-275 basepairs (bp)
- Primer size: min 15, opt 20, max 36 bp
- Primer melting temperature: min 50°C, optimal 60°C, max 65°C
- GC% content: min 40%, optimal 50%, high 60%
- Concentration of divalent cations = 3.8 mM
- Concentration of dNTPs needs to be equal to 0.8 mM
- Internal Oligo: size min 15, optimal 20, max 30 bp
- Internal Oligo: Melting temp min 62°C, optimal 63°C, max 70°C
- Internal Oligo: GC% min 30%, optimal 50%, max 80%

Table A2. The mean and standard deviation of the total number of copies of target per partition for EVD68 and PMMoV. The number of samples out of a random 36 included in this analysis that had detectable target in them is provided. Only these samples with detectable concentrations were used to calculated mean and standard deviation in the table.

| Target | EVD68 | PMMoV |
|---|---|---|
| Number of samples with detectable target (%) | 19 (53%) | 36 (100%) |
| Mean target copies per partition | $1.04 \times 10^{-3}$ | 0.150 |
| Standard deviation copies per partition | $1.18 \times 10^{-3}$ | 0.069 |
